# Eculizumab for the acute attack of neuromyelitis optica spectrum disorder

**DOI:** 10.1101/2025.06.17.25329603

**Authors:** Qingqing Zhuang, Wenjuan Huang, Jingzi ZhangBao, Zhouzhou Wang, Lei Zhou, Xiang Li, Yuxin Fan, Liang Wang, Yiqin Xiao, Jian Yu, Min Wang, Hongmei Tan, Chao Quan

## Abstract

**Objective:** To assess clinical outcomes in patients treated with eculizumab for acute attacks of aquaporin-4 antibody-positive neuromyelitis optica spectrum disorder (NMOSD).

**Methods:** We retrospectively analyzed prospectively collected data from the Huashan NMOSD registry cohort, and included patients who received eculizumab within 30 days of attack onset. Eculizumab was administered at 900 mg weekly for four weeks, followed by an eight-week observation period. Primary outcomes included visual acuity and visual field for optic neuritis (ON) and muscle strength, assessed using the Medical Research Council (MRC) scale for longitudinally extensive transverse myelitis (LETM). Serum neurofilament light chain (sNfL) and glial fibrillary acidic protein (sGFAP) levels were also monitored.

**Results:** Nine patients (seven with ON, two with LETM) were included. All patients received high-dose intravenous methylprednisolone prior to eculizumab treatment. Following eculizumab, six of the seven ON patients showed significant improvements in visual acuity and visual fields, with five recovering to near-normal or pre-attack vision. Visual field mean deviation improved from −22.4 dB to −2.0 dB (p = 0.008). Among LETM patients, one regained substantial lower limb strength (MRC grade 0 to 3 proximally, 4 distally), while the other showed improvement in distal strength (MRC grade 0 to 3). Serum sGFAP decreased from 278.0 to 130 pg/mL (p = 0.027), while sNfL levels transiently increased before stabilizing. LETM patients developed urinary tract infections, and one had Klebsiella pneumoniae pneumonia; all infections were effectively treated.

**Conclusion:** Eculizumab may yield favorable outcomes in acute NMOSD attacks, with infection monitoring being particularly important in severe cases.

## Introduction

Neuromyelitis optica spectrum disorder (NMOSD) is a rare autoimmune inflammatory demyelinating disease of the central nervous system (CNS), mediated by anti-aquaporin-4 immunoglobulin G (AQP4-IgG) antibodies (1). NMOSD is characterized by recurrent attacks of optic neuritis (ON) and longitudinally extensive transverse myelitis (LETM), frequently resulting in permanent vision loss and motor paralysis (2). Current therapeutic interventions for acute attacks primarily include high-dose intravenous methylprednisolone pulse therapy (IVMP) and therapeutic plasma exchange (TPE) (3). However, clinical outcome data indicate that only approximately 21.6% of patients attain complete recovery post-relapse, while 6% show no improvement at all (4). These findings underscore the critical need for therapies capable of rapidly attenuating acute tissue injury.

Complement-mediated enhancement of local inflammation and cytotoxicity are key elements in the pathophysiology of NMOSD. Upon binding to extracellular epitopes of AQP4, AQP4-IgG activates the complement system (5, 6) and ultimately forming C5b-9 membrane attack complex (MAC), driving astrocyte lysis, blood-brain barrier disruption and neuronal injury (7). Histopathological analyses have revealed that the sites of AQP4 loss within lesions correspond to regions of immunoglobulin and complement activation (8, 9). In the cerebrospinal fluid (CSF) of untreated NMOSD patients during the acute phase, increased levels of C5a and C5b-9 have been observed (10, 11).

Eculizumab, a humanized monoclonal antibody inhibiting complement C5, exerts its therapeutic effect by preventing the generation of C5a and MAC formation, thereby abrogating the pathological consequences of complement cascade activation (12). The phase III PREVENT trial (NCT01892345) established eculizumab as a first-line therapy for relapse prevention in AQP4-IgG-positive NMOSD (13). Rationales also exist for using eculizumab as an acute therapy for NMOSD relapse, as its rapid onset of action and continuous near-complete inhibition of C5 activity upon the first infusion (14).

Eculizumab has been approved for NMOSD in China since October 2023. The current study included nine patients with AQP4-IgG-positive NMOSD who were treated with eculizumab during the acute relapse phase. Changes in vision and motor functions, serological biomarkers, and retina measurements by optical coherence tomography (OCT) were assessed over an eight-week observation period.

## Methods

### Study design and participants

This is a retrospective analysis of prospectively collected data from the NMOSD cohort at Huashan Hospital. The study involved patients who were part of the cohort followed prospectively from their NMOSD diagnosis. Between October 2023 (when eculizumab was approved for NMOSD in China) and February 2025, 26 consecutive patients with AQP4-IgG-positive NMOSD received eculizumab, of whom 9 were included in this analysis. The inclusion criteria were as follows: (1) age ≥ 18 years; (2) diagnosed with AQP4-IgG-positive NMOSD according to the 2015 revised criteria (15); (3) received eculizumab during an acute phase of NMOSD, defined as within 30 days of attack onset, and (4) adherence to an 8-week follow-up from eculizumab initiation.

The decision to initiate eculizumab “early” was based on the aim of preventing complement-mediated neuronal damage and alleviating neurological deficits. This decision was made collaboratively between patients and physicians. As eculizumab is not reimbursed by national insurance, the decision was also influenced by the patients’ financial capacity.

Eculizumab was administered at a dosage of 900 mg weekly for four consecutive weeks. Since the drug is not covered by China’s national insurance, patients autonomously decided whether to continue eculizumab after receiving the four doses. Patients were advised to receive meningococcal vaccination at the earliest opportunity.

For those who were vaccinated, oral antibiotics were continued for two weeks following vaccination completion. In patients who did not receive vaccination, oral antibiotic prophylaxis was maintained for two weeks after the completion of eculizumab treatment.

Attacks were defined as followings and confirmed by a neurologist: (1) new neurological symptoms related to NMOSD lasting longer than 24 hours and occurring > 30 days after the previous attack; (2) no other underlying aetiology (e.g., fever or infection); (3) accompanying objective change in neurological examination, such as decreased muscle strength for myelitis and reduction in visual acuity or visual field defect for ON.

### Data collection

Data were extracted from the Huashan NMOSD prospective registry cohort, which includes regular follow-ups every six months during the remission phase, along with more intensive follow-ups during acute relapses. Demographic data and disease history prior to eculizumab treatment were collected, including age, gender, disease duration from the initial attack to the time of study inclusion, history of previous relapses, comorbidities, AQP4-IgG titers, and use of medications prior to the current attack. The time from the onset of the current attack to the initiation of IVMP, as well as the time from current attack onset to the initiation of eculizumab, were also recorded.

Clinical, serological, and ophthalmologic assessments were conducted for acute relapse treated with eculizumab, according to the protocol predefined in the NMOSD hospital registry. Specifically, assessments were conducted on the day of eculizumab initiation (W0), followed by weekly assessments for the first four weeks (Weeks 1, 2, 3, and 4) and again at Week 8 after eculizumab initiation. Each evaluation included vital signs, adverse event, the Expanded Disability Status Scale (EDSS) score, muscle strength in patients with myelitis attack, best corrected visual acuity (BCVA), visual fields, Visual Functional System Score (VFSS), and OCT measurements including peripapillary retinal nerve fiber layer (pRNFL) and macular ganglion cell-inner plexiform layer (mGCIPL) in patients with ON attack. Serum neurofilament light chain (sNfL), serum glial fibrillary acidic protein (sGFAP), complete blood count, C-reactive protein, and hepatorenal functions were determined on the day of eculizumab initiation (W0) and at Weeks 0, 2, 4 and 8.

### Outcome Measures

The primary outcome was the change in disability status, including muscle strength measured by the Medical Research Council (MRC) scale for patients with LETM, BCVA and visual field for patients with ON. The secondary outcome was EDSS, VFSS, and adverse event. The exploratory outcomes were changes of serological biomarkers (sGFAP and sNfL) and OCT measurements (pRNFL and mGCIPL).

### AQP4-IgG

AQP4-IgG detection was conducted using a fixed-cell-based assay with indirect immunofluorescence, utilizing HEK-293 cells transfected with M1-AQP4 at Huashan Hospital.

### sGFAP and sNfL

The collected serum samples were immediately separated by centrifugation and frozen at −80 °C until analysis. sGFAP and sNfL concentrations were measured in duplicates via ultra-sensitive single-molecule arrays (SIMOA). Neurology 2-plex B kits were used with an HD-1 immunoassay analyzer (Quanterix, Boston, Massachusetts, USA) as per the manufacturer’s instructions.

### BCVA and Visual field

BCVA was assessed using the Standard Logarithmic Visual Acuity Chart (GB/T 11533-2011) in accordance with Chinese national standard(16), and a BCVA below 0.01 was documented with count fingers (CF) or hand movements (HM).

The central visual field was assessed using a Humphrey visual field analyzer (Carl Zeiss, Germany). Swedish Interactive Thresholding Algorithm strategy 30-2, No. III white visual label was selected as the detection program, and the visual field mean deviation (MD) was automatically calculated. The reliability criteria were false-positive and false-negative rates of < 33% and fixation losses of < 20%.

### OCT

TOPCON’s sweep OCT Triton (Topcon Healthcare, Tokyo, Japan) was used with a speed of 100000 A-scans per second. The peripapillary pRNFL thickness was measured using the “3D Disc 6.0*6.0mm” mode, which scans a circular area with a 6-mm diameter centered on the optic disc. The ganglion cell complex thickness was acquired using the “3D Macula 7.0*7.0mm” mode, which captures data from a square grid (7 mm × 7 mm) centered on the macula. The device automatically segmented the measurements into three components: macular retinal nerve fiber layer (mRNFL), mGCIPL, and mGCIPL plus mRNFL. Only high-quality images with signal strength index ≥ 40 were accepted according to the OSCAR-IB criteria(17).

### Statistical Analysis

Quantitative data are presented as median and range. Categorical variables were described by counts and percentages. The comparison between pre- and post-treatment was conducted using the paired Wilcoxon signed-rank test. All statistical analyses were performed using R version 4.4.1(R Foundation). Graphs were generated using R version 4.4.1 and adjusted using Adobe Illustrator version 27.0 (Adobe Inc.). The level of statistical significance was set at *p* < 0.05.

## Result

### Patient characteristics

This study included nine patients diagnosed with AQP4-IgG-positive NMOSD, comprising seven females and two males. Among them, four patients experienced their first NMOSD attack, while the remaining had a history of 1 to 9 previous attacks (Table 1).

**Table 1.** Demographic and clinical assessments before and after eculizumab.

| Patient No. | Sex | Age of attack (years) | Disease duration<sup>†</sup> (months) | Attack type | EDSS before attack | BCVA before attack (L, R) | No. of previous attacks | Pervious therapy | Comorbidities | Serum AQP4-Ig G titer | Attack onset to IVMP (days) | Attack onset to Ecu (days) | EDSS before Ecu | EDSS after Ecu (W8) | BCVA before Ecu (L, R) | BCVA after Ecu (W8) (L, R) | Other acute therapies |
| --- | --- | --- | --- | --- | --- | --- | --- | --- | --- | --- | --- | --- | --- | --- | --- | --- | --- |
| 1 | F | 30-34 | 24 | TM | 0 | - | 1 | MMF | SS, SLE | 1:32 | 1 | 13 | 8 | 7.5 | - | - | IVMP, TPE |
| 2 | F | 40-44 | 24 | LON | 3 | 1.0, 0.12 | 1 | MMF | ANFH, SLE | 1:320 | 2 | 4 | 3 | 3 | 0.8, 0.1 | 1.0, 0.12 | IVMP, IVIg |
| 3 | F | 35-39 | 0.4 | LON | 0 | 1.0, 1.0 | 0 | - | SS | 1:320 | 5 | 13 | 2 | 1 | 1.0, 1.0 | 1.0, 1.0 | IVMP |
| 4 | M | 35-39 | 25 | LON | 0 | 1.0, 1.0 | 1 | - | - | 1:100 | 9 | 17 | 3 | 1 | CF/20cm, 1.0 | 0.9, 1.0 | IVMP |
| 5 | F | 65-69 | 148 | LON | 6 | 0.5, HM/10cm | 9 | Satr | - | 1:1000 | 1 | 3 | 6 | 6 | 0.05, HM/10cm | 0.5, HM/10cm | IVMP |
| 6 | M | 65-69 | 0.13 | TM | 0 | - | 0 | - | TTX | 1:100 | 4 | 5 | 8 | 8 | - | - | IVMP |
| 7 | F | 25-29 | 2 | BON | 0 | 1.0, 1.0 | 3 | - | - | 1:32 | 3 | 7 | 4 | 1 | CF/30cm, 0.08 | 1.0, 1.0 | IVMP |
| 8 | F | 35-39 | 0.7 | RON | 0 | 1.0, 1.0 | 0 | - | - | 1:320 | 7 | 21 | 3 | 1 | 1.0, CF/30cm | 1.0, 1.0- | IVMP |
| 9 | F | 25-29 | 0.3 | LON | 0 | 1.0, 1.0 | 0 | - | - | 1:1000 | 9 | 10 | 3 | 3 | HM/10cm, 1.0 | HM/10cm, 1.0 | IVMP×2<sup>‡</sup> |

<sup>‡</sup>Patient received two courses of IVMP.
<sup>†</sup>Duration refers to the time from the first attack to the current admission to Huashan Hospital
ANFH, Avascular Necrosis of the Femoral Head; BCVA, best corrected visual acuity; ON, optic neuritis; BON, bilateral ON; Ecu, eculizumab; EDSS, Expanded Disability Status Scale; Inebi, inebilizumab; IVIg, intravenous immunoglobulin; IVMP, intravenous methylprednisolone pulse; LON, left ON; MMF, Mycophenolate mofetil; RON, right ON; Satr, satralizumab; SLE, Systemic Lupus Erythematosus; SS, Sjögren's syndrome; TM, transvers myelitis; TPE, therapeutic plasma exchange; TTX, Thyrotoxicosis

Two patients (Patients 1 and 6) presented with LETM. Patient 1 had spinal cord lesions extending from C3 to T7, whereas Patient 6 had involvement from C1 to T2. The T2 sequence of magnetic resonance imaging (MRI) in both patients revealed hyperintense spinal cord signals accompanied by cord swelling.

Eight eyes from seven patients (Patients 2–5 and 7–9) were affected by ON. During the follow-up, ophthalmological examination data were missing for Patients 2 and 9 at Week 4. Orbital MRI was performed on five of these seven patients, revealing abnormal optic nerve signals with enhancement. Patient 7 exhibited bilateral involvement, while the other patients had unilateral ON.

Among all nine patients, the median nadir EDSS score prior to eculizumab treatment was 3 (range 2–8). Of the eight affected eyes, six exhibited a nadir BCVA below 0.1.

All nine patients received IVMP with an initial dose of 1000 mg, with two also undergoing TPE (Patient 1) or intravenous immunoglobulin (IVIg, Patient 2) before starting eculizumab (Figure 1F). Patient 9 received two rounds of IVMP because of unsatisfactory visual recovery. The median time from symptom onset to eculizumab initiation was 10 (range 3–21) days.

**Figure 1.**
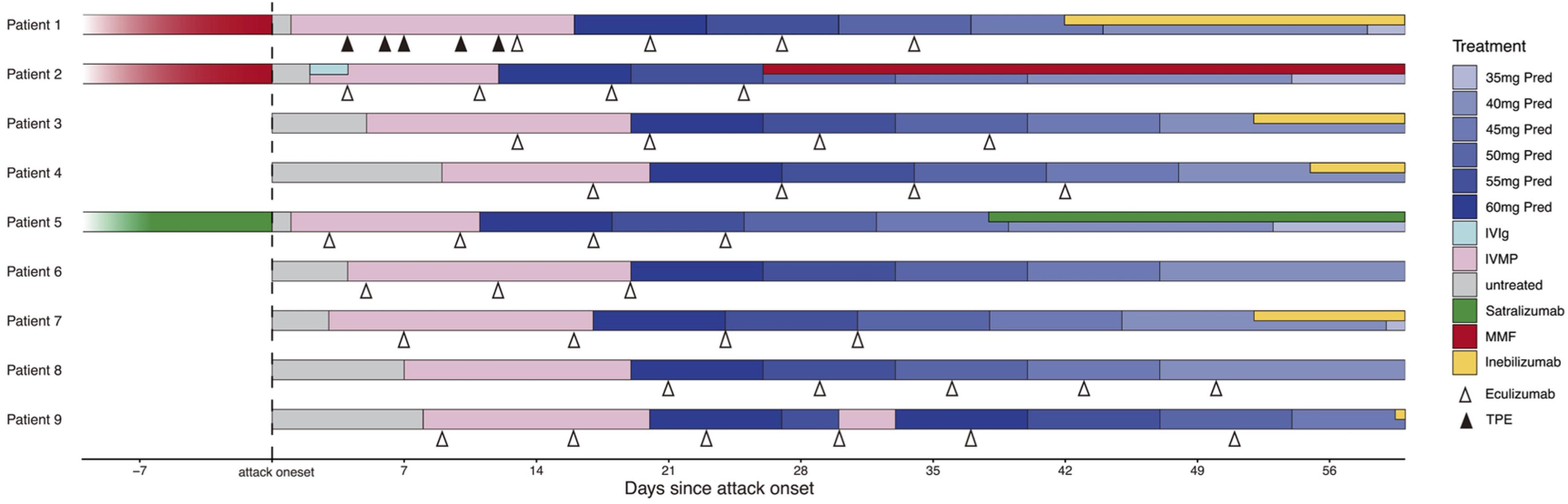
Treatment of the nine patients before and during NMOSD acute attack. Pred, prednisone; IVIg, intravenous immunoglobulin; IVMP, intravenous methylprednisolone pulse; MMF, Mycophenolate mofetil; TPE, therapeutic plasma exchange.

### Improvement in neurological function

Eight out of nine patients completed the full course of four weekly doses of eculizumab, while one patient (Patient 6) discontinued treatment after receiving three doses. Disability improved following eculizumab treatment, as shown in Figure 2. The median EDSS score decreased from 3 (range 2–8) before eculizumab treatment to 3 (range 1–7.5) at week-8 of follow-up (*p* = 0.042).

**Figure 2.**
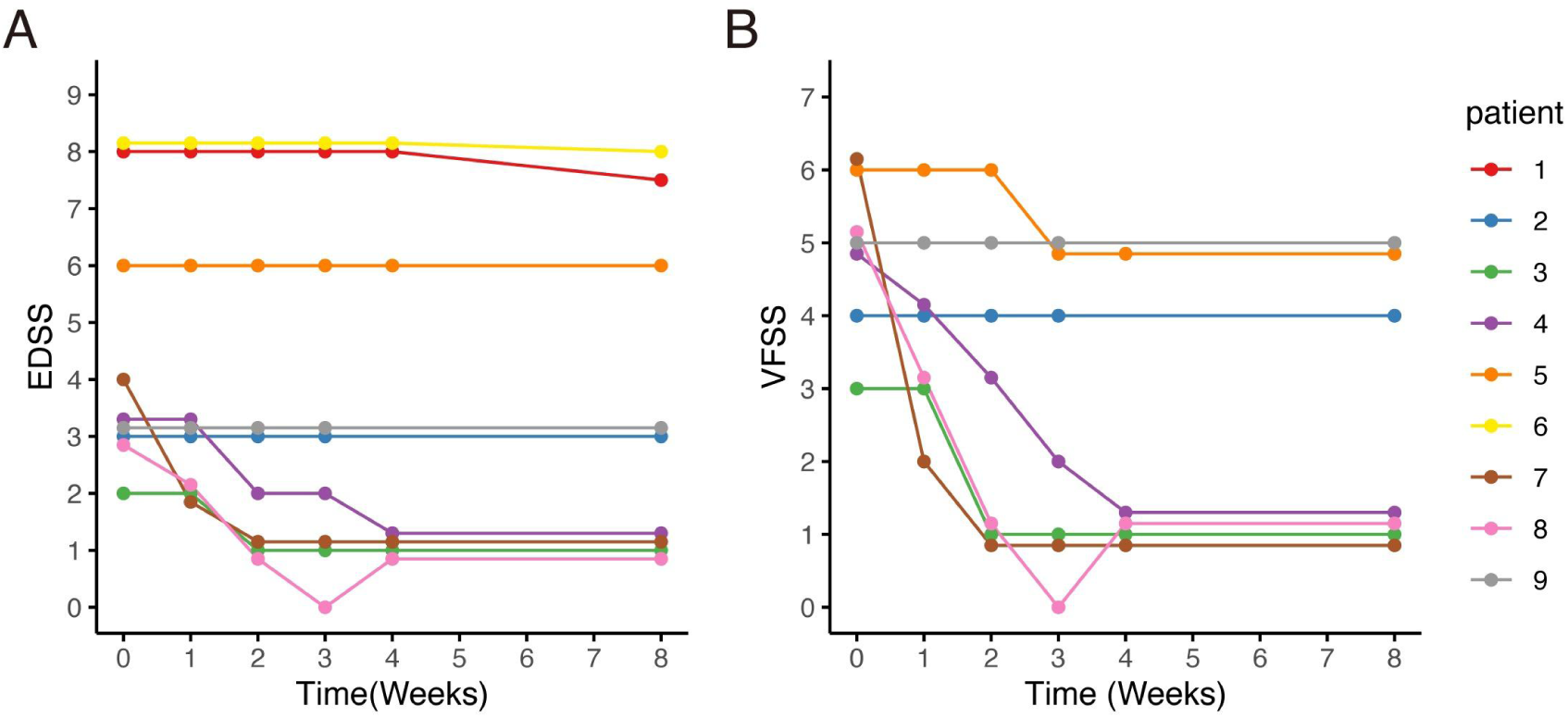
EDSS score, and VFSS changes after eculizumab initiation. (A, B) After eculizumab initiation, both the EDSS and VFSS scores demonstrated either improvement or stability. (B) Among the 7 patients with optic neuritis, Patient 2 had severe visual field defects and a BCVA of 0.12 in the contralateral eye due to a previous optic neuritis, while Patient 5 had only HM/10cm in the contralateral eye due to a previous optic neuritis. Consequently, no obvious changes were observed in the VFSS for the two patients. Patient 9 did not experience visual improvement, while the VFSS of the remaining patients decreased to 1. BCVA, best corrected visual acuity; EDSS, Expanded Disability Status Scale; HM: hand movements; VFSS, Visual Functional System Score.

#### Myelitis

Patient 1 (30-34 years, female) with LETM, began IVMP therapy one day after symptom onset, followed by five sessions of TPE. However, her lower extremity muscle strength remained at grade 0 after the final TPE session. Eculizumab therapy was initiated 13 days after symptom onset, leading to a gradual recovery of muscle strength. She received a total of four weekly doses of eculizumab, and by week 8, her lower extremity strength had improved to grade 3 proximally and grade 4 distally (Table 2). She was subsequently transitioned to the inebilizumab regimen. During a follow-up visit six months after the final dose of eculizumab, despite being beyond the observation window of this study, the patient was able to walk 500 meters independently.

**Table 2.**
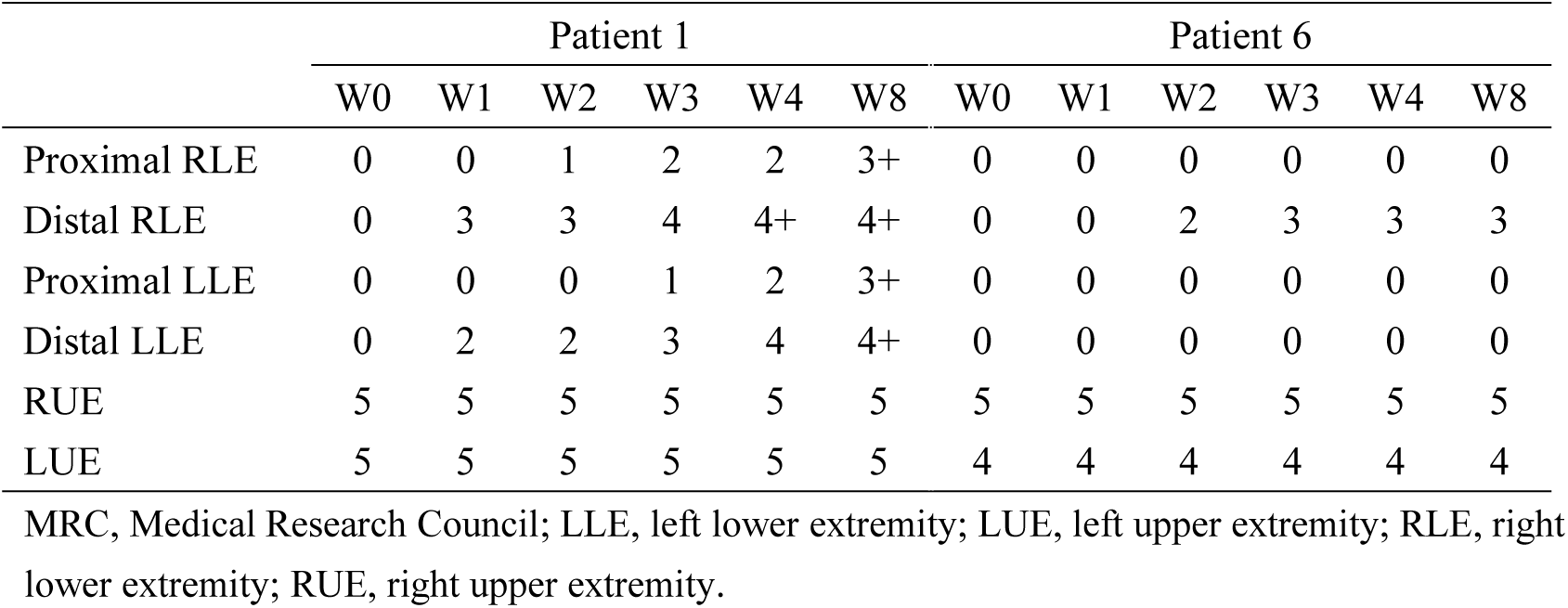
Muscle strength changes by MRC scale score of Patient 1 and 6 after eculizumab treatment initiation.

Patient 6, (65-69 years, male) , developed ascending myelitis involving nearly the entire cervical cord. He experienced complete paralysis of the lower limbs, left upper limb weakness, and a gradual ascension of the sensory level. His baseline sGFAP level was markedly elevated at 26,360.0 pg/mL (Figure 3A). IVMP therapy was initiated four days after symptom onset, followed by eculizumab on the second day after IVMP initiation. After starting eculizumab, no further progression of disability was observed. By week 2 of treatment, he regained movement in the distal right lower limb, coinciding with a sharp decline in sGFAP levels to 292.0 pg/mL. He continued weekly eculizumab treatments for three weeks. However, despite prophylactic antibiotic coverage, he developed a *Klebsiella pneumoniae* pneumonia, leading to the discontinuation of eculizumab therapy. Intravenous antibiotics were initiated and effectively controlled the infection. By week 8, muscle strength in his distal right extremities stabilized at grade 3 (Table 2).

**Figure 3.**
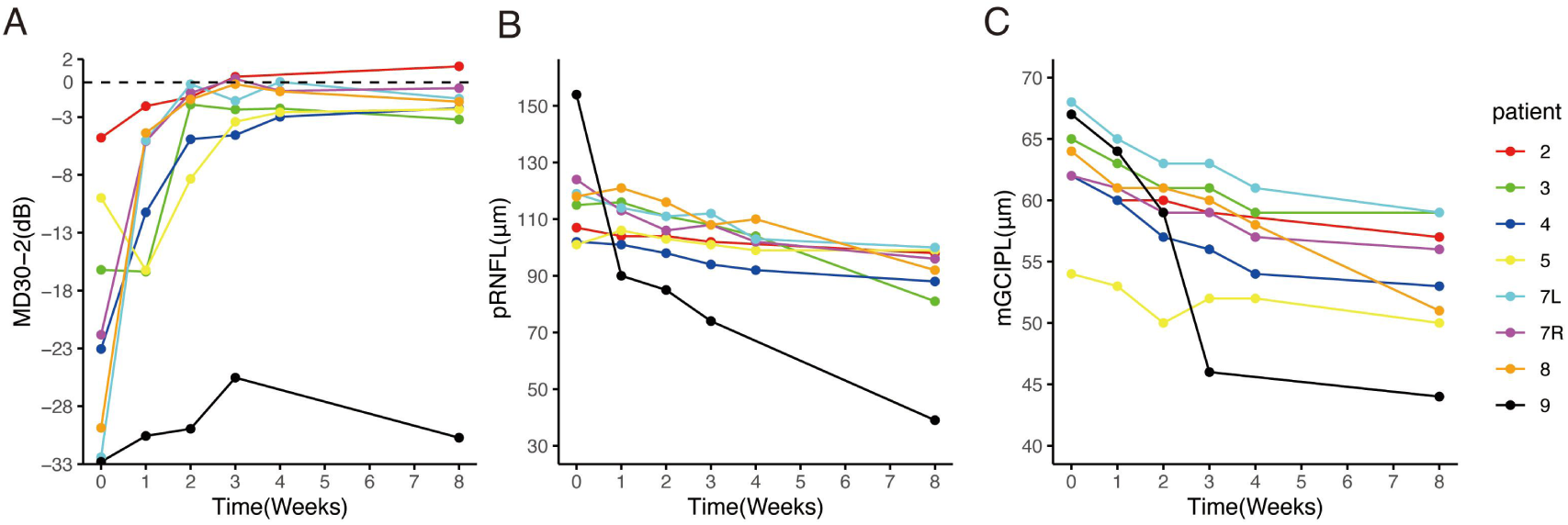
Changes in sGFAP and sNfL levels of each patient after eculizumab initiation. (A, C) The median sGFAP level was 278.0 pg/ml before eculizumab treatment and decreased to 130.0 pg/ml after 2 weeks of treatment (*p* = 0.027). Then the sGFAP levels tended to be stable, and there was no significant difference between Week 2 and Week 4, and week 4 and week 8. (B, D) The middle horizontal line represents the median, and the box represents the interquartile range. The sNfL levels showed an upward trend after the initiation of eculizumab treatment, reaching its peak at Week 4, then slightly decreased at Week 8, yet there was no statistically significant difference between each week. sGFAP, serum glial fibrillary acidic protein; sNfL, serum neurofilament light chain.

#### ON

Seven patients received eculizumab treatment during an ON attack, affecting eight eyes. Among them, four patients (Patients 2, 3, 7, and 8) demonstrated prominent visual recovery in five eyes, with BCVA improving to 1.0 and visual fields nearly returning to normal by Week 8. Notably, Patient 5, who had a history of recurrent ON attacks, also experienced restoration of both BCVA and the visual field in her left eye to baseline levels prior to this attack (Figure 4). In Patient 4, BCVA improved to 0.9, though some residual visual field defects persisted. Altogether, among the 8 affected eyes of the seven patients, the median visual field MD significantly improved from −22.4 dB (range −32.8 to −4.8) to −2.0 dB (range −30.7 to 1.4) (*p* = 0.008) (Figure 5A). The VFSS decreased from 5 (range 3 to 6) to 1 (rang 1 to 5) after treatment (*p* = 0.042).

**Figure 4.**
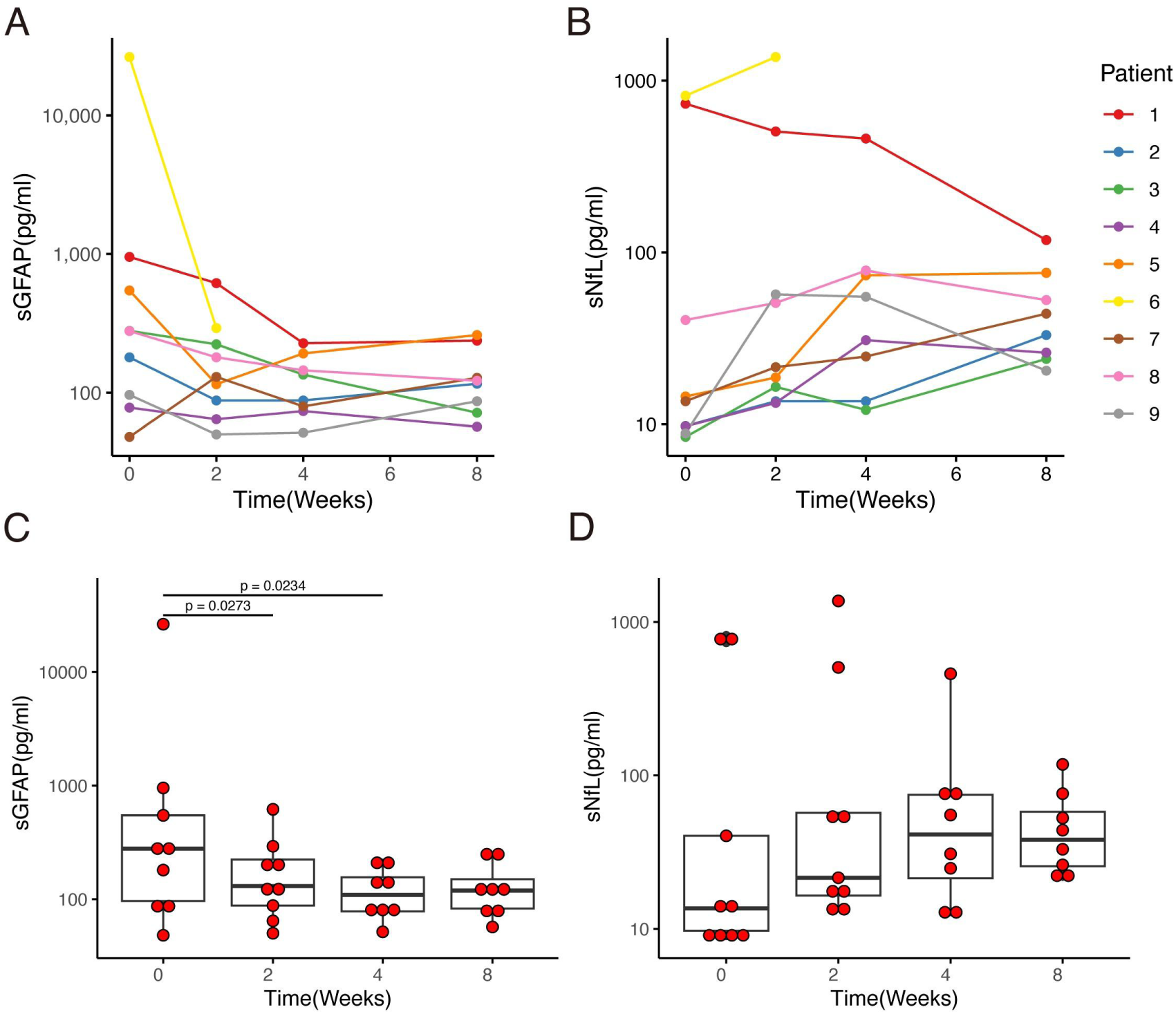
Changes of visual fields and BCVA in the seven patients with optic neuritis attack treated with eculizumab. The visual fields are primarily depicted using pattern deviation. For visual fields with severe depression where pattern deviation cannot be reliably calculated, total deviation is shown instead (for Patient 4, Patient 7L, Patient 7R, Patient 8 at W0, and Patient 9 at all time points). BCVA, best corrected visual acuity; CF, count finger; HM, hand movement; L, left eye; NA, not performed; R, right eye.

**Figure 5.**
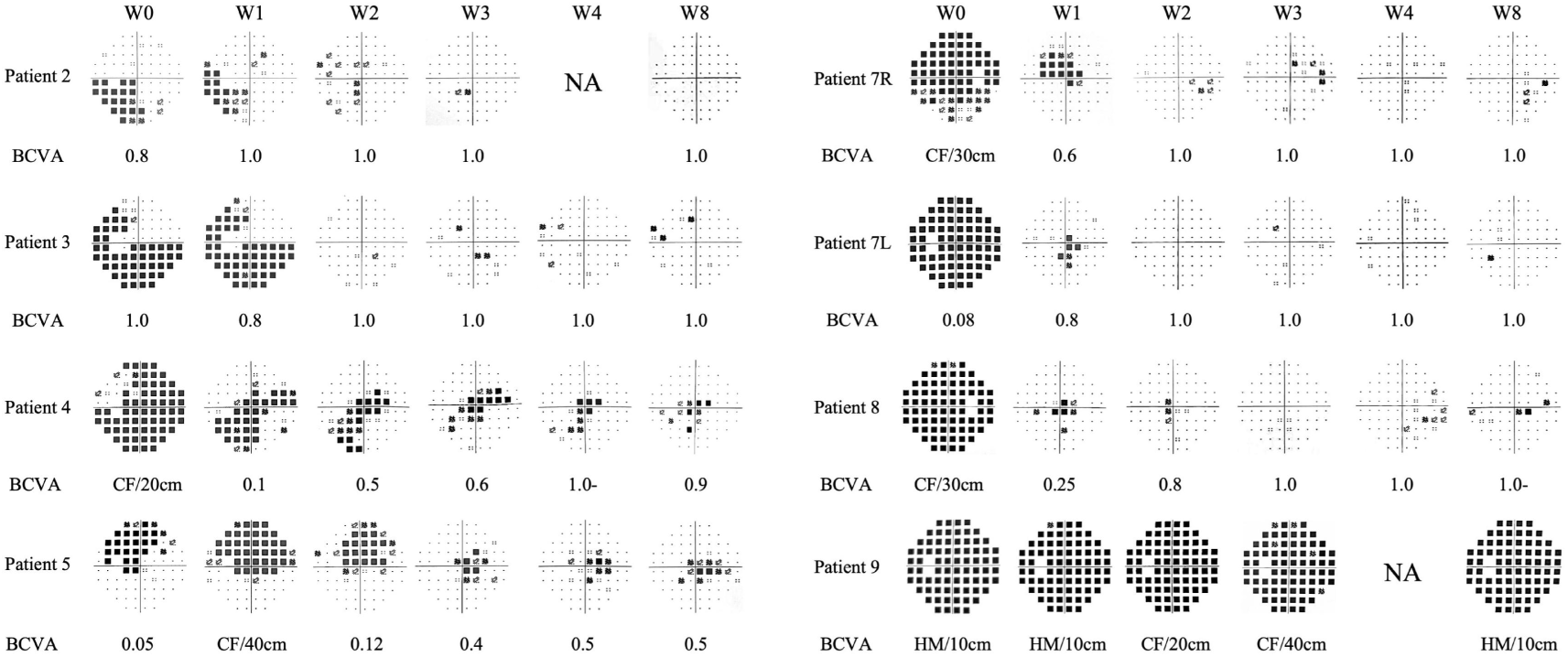
Change of visual field MD30-2, pRNFL and mGCIPL after eculizumab treatment initiation. (A) The MDs of the 8 eyes of the 7 patients with ON attack improved prominently after eculizumab treatment initiation; (B, C) the thickness of pRNFL and mGCIPL decreased over the 8-week observation period. Patient 9 exhibited faster pRNFL and mGCIPL atrophy compared to other patients. MD, mean deviation; mGCIPL, macular ganglion cell-inner plexiform layer; pRNFL, peripapillary retinal nerve fiber layer.

Patient 9 was the only patient whose visual function did not recover. She began IVMP therapy on day 9 after symptom onset and started eculizumab on day 10. Prior to this, she underwent extensive diagnostic evaluations, including waiting for optic nerve MRI and AQP4 antibody results, during which her visual field continued to deteriorate (Supplementary Figure 1) and her visual acuity decreased to HM. IVMP was only initiated after she presented to our center. Her serum AQP4-IgG titer was very high (>1:1000), and MRI revealed a strikingly long-segment enhancement of the left optic nerve in the intraorbital region, with pronounced nerve thickening and tortuosity (Supplementary Figure 1). Her baseline pRNFL thickness was also greater than that of other patients. Despite two rounds of IVMP and eculizumab therapy, her visual function did not improve at week 8, and the improvement in MD was also unsatisfactory (Figure 4).

### pRNFL and mGCIPL alteration

Through the initial weekly OCT examinations and a final examination on week 8, it was observed that despite all seven patients with an ON attack receiving IVMP and eculizumab treatment, both the mGCIPL and pRNFL exhibited progressive thinning over the course of eight weeks (Figure 5B, C). In Patient 9, who showed poor visual recovery, a more rapid progression of retinal thinning was observed, with the rate faster than that of the other patients (Figure 5B, C). Compared to other patients, Patient 8 exhibited a faster rate of retinal atrophy during weeks 4–8. Correspondingly, we observed a slight decline in her vision and visual field at week 8 compared to week 4.

### sGFAP and sNfL

Baseline sGFAP and sNfL levels in the two patients with acute myelitis were remarkably higher than those with ON. In the patients with acute myelitis, sGFAP levels were measured at 26,360.0 pg/ml (Patient 6) and 952.0 pg/ml (Patient 1), while sNfL levels were 816.0 pg/ml (Patient 6) and 733.0 pg/ml (Patient 1). The seven patients with ON exhibited a median sGFAP level of 180.0 (range 48.0–546.0) pg/ml and a median sNfL level of 9.8 (range 8.4–40.4) pg/ml.

Following eculizumab treatment, sGFAP levels showed a notable decline within four weeks. Statistically significant reductions were observed between Week 0 and Week 2 (median 278.0 vs 130.0 pg/ml, *p* = 0.027) and between Week 0 and Week 4 (median 278.0 vs 111.4 pg/ml, *p* = 0.023). However, no significant differences were noted between Week 2 and Week 4 (median 130.0 vs 111.4 pg/ml, *p* = 0.311), or Week 4 and Week 8 (median 111.4 vs 119.0 pg/ml, *p* = 0.461) (Figure 3A, C).

Conversely, sNfL levels exhibited an upward trend, peaking at Week 4 of eculizumab treatment and then stabilized. Despite this increase, no statistically significant differences were detected across various time points (Figure 3B, D).

### Safety

At the initiation of eculizumab therapy, oral antibiotic prophylaxis was administered to all nine patients. Specifically, Patients 1–5 and 7, who tested negative on penicillin skin testing, received phenoxymethylpenicillin potassium tablets. Patients 6 and 8, with positive penicillin skin tests, were prescribed oral cephalosporin antibiotics. Patient 9, with a documented cephalosporin allergy, was administered oral levofloxacin. Meningococcal vaccination was administered to four patients (Patients 1, 3-5) within two weeks following the first dose of eculizumab. The remaining four unvaccinated patients continued oral penicillin prophylaxis for two weeks after the completion of eculizumab therapy. No cases of meningococcal infection were observed during the study period.

Patient 1, who had spinal cord involvement and required urinary catheterization, developed a urinary tract infection during the fourth week of eculizumab therapy. Patient 6, who experienced a myelitis flare, developed *Klebsiella pneumoniae* pneumonia during the third week of eculizumab treatment and subsequently discontinued therapy. Following appropriate intravenous anti-biotics, the infections in both patients were effectively controlled. No adverse reactions were observed in patients with ON, and their vital signs, routine blood test results, and liver and renal function parameters remained within normal limits. None of the patients experienced hypertension, headache, diarrhea, nausea and vomiting, skin rash, anemia, or thrombocytopenia.

### Maintenance therapy after eculizumab

Following acute treatment with eculizumab, five patients switched to inebilizumab as their maintenance therapy, while one patient received satralizumab, and another was treated with mycophenolate mofetil. Only one patient (Patient 8) continued eculizumab biweekly for recurrence prevention. One patient did not receive any additional preventive treatment aside from oral steroids (Figure 1).

## Discussion

Solid evidences support eculizumab as a preventive therapy for NMOSD (13, 18); however, its potential role in treating acute attacks of NMOSD remains underexplored. Since complement activation plays a key role in driving neuroinflammation and tissue damage during NMOSD attacks, and eculizumab rapidly lowers free C5 concentrations below the threshold required for complete terminal complement inhibition within just 60 minutes of infusion (19), it is anticipated that eculizumab could also be beneficial in the acute phase of NMOSD.

To date, 13 cases have been reported in which eculizumab was administered early after the onset of an NMOSD attack, demonstrating its potential in mitigating the worsening of neurological disability associated with these attacks (Supplementary table 1) (20–26). However, among these cases, only six received eculizumab within 30 days of attack onset. Moreover, key supportive evidence of effectiveness, such as dynamic changes in visual fields and retinal thickness, or neuroinjury-related biomarkers like sGFAP and sNfL were not investigated in these cases, leaving critical gaps in our understanding of eculizumab’s therapeutic potential in the acute phase.

In the current study, nine patients receiving eculizumab after 10 (range 3-21) days from the onset of acute attacks were analyzed. Seven with ON and two with myelitis attack. Over the subsequent 8-week observation period, the majority of cases showed improvement in disability.

Seven patients experiencing an ON attack initiated eculizumab treatment between the 3rd and 21st day after onset. Six of them demonstrated remarkable recovery of visual function. Among these, five patients regained visual acuity ranging from 0.9 to 1.0, with their visual fields largely or nearly fully restored. Additionally, one patient with a history of multiple ON episodes recovered both BCVA and visual field to their pre-attack status.

The dynamic changes in the pRNFL and mGCIPL following acute ON remain incompletely understood. In this acute-phase study, we longitudinally monitored these changes over time. Despite all patients receiving eculizumab in combination with high-dose corticosteroid pulse therapy, a progressive trend of retinal atrophy was still observed. Notably, in the only patient with poor visual recovery (Patient 9), both the rate and extent of retinal atrophy were more pronounced. This suggests that rapid and substantial retinal atrophy during the acute phase correlates with unfavorable visual outcomes. Furthermore, it indicates that once inflammation is triggered, secondary tissue degeneration appears to be inevitable, regardless of the intensity of therapeutic intervention. The current 8-week observation period may be insufficient to determine when retinal atrophy stabilizes (27). Future studies are warranted to further investigate this issue.

Patient 9, who exhibited poor visual recovery, highlights the necessity of timely intervention. Corticosteroid therapy was initiated on day 9 post-symptom onset, followed by eculizumab on day 10. Delayed treatment due to prolonged waiting for optic nerve MRI and AQP4-IgG results allowed progressive visual field deterioration, culminating in complete visual field loss upon arrival at our center. Her markedly elevated AQP4-IgG titer (>1:1000), pronounced optic nerve enhancement and swelling on MRI, and pRNFL thickening indicated severe inflammatory activity. We speculate that such aggressive inflammation has a critically narrow therapeutic window. Despite two courses of 1000 mg IVMP and eculizumab therapy, visual recovery remained poor, emphasizing the imperative for early intervention to improve outcomes.

Following treatment initiation, patients with myelitis exhibited varying degrees of muscle strength improvement. Patient 1 showed a reduction in EDSS score from 8 to 7.5. Patient 6 achieved disease stabilization, with no further sensory level ascension. Although the overall improvement in Patient 6 was modest, it is noteworthy that this elderly patient presented with potentially life-threatening LETM. The attainment of a favorable trajectory within the 8-week observation period is therefore clinically meaningful.

The speed of recovery from myelitis within the 8-week period appeared slower than that observed in ON, likely due to the greater lesion burden. However, beyond the initial phase of recovery—primarily driven by the resolution of inflammation and edema—it is anticipated that a prolonged period of neurological repair will occur over 6–24 months. During this phase, the optimization of alternative neural circuits through physical and occupational therapy may facilitate further functional improvement (28). Therefore, an 8-week observation period may be insufficient to fully assess recovery following acute myelitis. At six months post-attack, Patient 1 regained the ability to ambulate independently for 500 meters, although the effects of steroids and subsequent inebilizumab treatment cannot be excluded. However, Patient 6 was lost to follow-up.

Through this study, we were able to characterize the dynamics of sGFAP and sNfL following an acute attack. A distinct pattern emerged: sGFAP levels demonstrated a significant decline within the first two weeks of eculizumab treatment, after which the rate of decline slowed. In contrast, sNfL levels showed a gradual upward trend despite eculizumab treatment, suggesting the occurrence of delayed neuro-axonal damage secondary to astrocytopathy. This finding aligns with a recent report indicating that sGFAP levels peak within the first week, whereas sNfL levels peak at five weeks post-attack (29) . However, due to the absence of a control group, we cannot confirm whether eculizumab attenuated the extent of the increase in sGFAP and sNfL levels.

No new safety concerns were identified in this cohort of acute attacks. This study demonstrated that eculizumab was generally well-tolerated for the treatment of NMOSD patients with acute ON. It is important to note that two patients with acute LETM developed urinary tract infections, and one patient developed a pulmonary infection. However, the potential association between these infections and NMOSD itself should also be considered, as severe myelitis is known to increase the risk of both pulmonary and urinary tract infections, especially in elderly patients. Notably, no cases of meningococcal infection were observed in this study.

In our cohort, only one patient underwent TPE prior to eculizumab administration, while the remainder received eculizumab alone. This pattern may reflect a clinical challenge: although TPE is supported by class I evidence particularly in corticosteroid-refractory myelitis (30), it presents logistical complexities when used alongside eculizumab. Specifically, TPE can remove circulating monoclonal antibodies such as eculizumab from plasma, necessitating a delay in eculizumab initiation if TPE is anticipated. Conversely, when TPE is required following eculizumab administration, re-dosing of eculizumab may be necessary to maintain therapeutic drug levels. Notably, substituting eculizumab for TPE may entail potential risk, particularly in severe or refractory cases where TPE has demonstrated benefit. Further comparative studies are needed to determine the optimal sequencing or combination of these therapies, given their potential pharmacokinetic interactions.

The primary limitation of this study is the absence of a control group, which prevents the exclusion of confounding factors, such as the therapeutic effects of other acute treatments (corticosteroids, TPE, and IVIg) and potential spontaneous improvement associated with the natural course of the disease. Additionally, selection bias may have been introduced by both physicians and patients due to the consent-driven treatment decision. Therefore, prospective randomized controlled trials are needed to rigorously assess the efficacy and safety of eculizumab as an acute treatment for AQP4-antibody-positive NMOSD attacks. If its benefit in the acute setting is definitively established in the future, early initiation of eculizumab could serve a dual purpose: aiding recovery from the current attack and preventing future relapses. Ravulizumab, which has recently demonstrated efficacy in AQP4-positive NMOSD (31), may also emerge as a promising candidate for acute intervention, pending confirmation of a sufficiently rapid onset of action.

## Conclusion

In conclusion, this study observed potentially favorable outcomes in patients treated with eculizumab during acute NMOSD attacks. However, careful monitoring for infection is warranted, particularly in elderly patients with severe myelitis.

## Data Availability

Data are available on reasonable request.

## Data availability statement

Data are available on reasonable request.

## Ethical standards

The current analysis (HIRB-KY2025-062), as well as the prospective NMOSD hospital registry (HIRB-KY2020-007) were approved by the Medical Ethics Committee of Huashan Hospital, respectively. Patients gave informed consent to participate in the studies before taking part.

## Author contributions

Dr Qingqing Zhuang participated this study during her subspecialty training in neuroimmunology at Huashan Hospital.

Qingqing Zhuang: data curation, formal analysis, investigation, methodology and writing (original draft, review and editing); Wenjuan Huang and Jingzi Zhangbao: data curation, formal analysis and writing (original draft, review and editing); Zhouzhou Wang and Lei Zhou: data curation and writing (original draft, review and editing); Xiang Li, Yuxin Fan, Liang Wang, Yiqin Xiao, Jian Yu, and Min Wang: writing (review and editing); Hongmei Tan: conceptualization, formal analysis, investigation; writing–review and editing, project administration, supervision; Chao Quan: conceptualization, formal analysis, investigation; writing–review and editing, funding acquisition, project administration, supervision, being responsible for the overall content as the guarantor

