## Supplemental Data 1 for "Eculizumab for the acute attack of neuromyelitis optica spectrum disorder"

Supplementary Materials

Table 1 Thirteen cases reported in which eculizumab was administered early after the onset of an NMOSD attack

| Reference | Age of attack (years)^a^ | Sex | Ethnics | Disease duration before the attack | Attack numbers before the attack | This attack type | Acute treatment and courses before Ecu | From attack onset to Ecu (days) | EDSS and VA (R, L) before attack | EDSS before Ecu treatment | VA before Ecu treatment (R, L) | EDSS after Ecu treatment | VA after Ecu treatment (R, L) | Prophylactic anti-infective treatment or meningococcal vaccination | Adverse event during Ecu treatment |
| --- | --- | --- | --- | --- | --- | --- | --- | --- | --- | --- | --- | --- | --- | --- | --- |
| Watanabe et al. (Case 1) | 50-54 | Female | NA | First attack | 0 | TM（C1-C7） | IVMP×2, TPE ×3 | 46 | EDSS 0  NA | 6.5 | NA | 4.5 | NA | vaccination+ceftriaxone | not detected |
| Watanabe et al. (Case 2) | 50-54 | Female | NA | 10 years | 2 | BS, TM | IVMP×2, TPE ×5 | 41 | EDSS 2.0  NA | 4.0 | 1.2, 1.2 | 3.5 | 1.2, 1.2 | vaccination+ceftriaxone | not detected |
| Watanabe et al. (Case 3) | 75-79 | Female | NA | 27 years | frequent | BON | IVMP×1, TPE ×3 | 30 | EDSS 7.5  0.3, 0.5 | 7.5 | 0.06, 0.05 | 7.5 | 0.4, 0.3 | vaccination+ceftriaxone | not detected |
| Watanabe et al. (Case 4) | 50-54 | Female | NA | 3 months | 1 | RON | IVMP×2, TPE×5, IVIg | 42 | EDSS NA  0.15, 1.2 | NA | 0.08, 1.2 | NA | 0.10, 1.2 | NA | not detected |
| Watanabe et al. (Case 5) | 90-94 | Male | NA | 5 years | 2 | BON | IVMP×1, TPE ×3 | 61 | EDSS 6.5  0.8, 0.9 | 6.5 | CF 20cm, 0.15 | 6.5 | 0.01, 0.2 | vaccination+ceftriaxone | not detected |
| Gorriz et al. | 45-49 | Female | Nigerian | 3 months | 1 | TM, BS, BON | IVMP, TPE | NA | EDSS 6.0  NA | 9.5 | NA | 7.5 | NA | vaccination+cephalosporins | Moderate anemia and thrombocytopenia |
| Chatterton et al. | 45-49 | Female | Chinese | 15 years | 3 | LON | IVMP,  TPE ×5 | 12 | EDSS NA  6/60, NA | NA | 6/60, LP | NA | 6/60, loss of central vision | vaccinations + Amoxicillin 250mg daily | NA |
| Enriquez et al. | 10-14 | Male | Ethiopia | First attack | 0 | BS | IVMP,  TPE ×2 | 6 | EDSS 0  1.2, 1.2 | 9.0 | 1.2, 1.2 | 2 | 1.2, 1.2 | vaccinations + antibiotic | left-hand tonic spasms |
| San-Galli et al. (Case 1) | 40-44 | Female | Afro-Guyanese | First attack | 0 | TM, BS | IVMP,  TPE ×10 | 32 | EDSS 0  1.2, 1.2 | 9.5 | NA | 8 | NA | vaccinations+oxacillin | NA |
| San-Galli et al. (Case 2) | 30-34 | Female | Ivorian | 2 years | 3 | BON, TM, BS | IVMP,  TPE ×10 | 21 | EDSS 8  NA | NA | NA | 4 | NA | vaccinations+oxacillin | NA |
| Kaneko et al. (Case 1) | 50-54 | Female | Japanese | 5 years | 3 | TM | IVMP,  TPE ×10 | 22 | EDSS 2  NA | 3.5 | NA | 2.5 | NA | vaccination | NA |
| Kaneko et al. (Case 2) | 50-54 | Female | Japanese | 20 years | 4 | BON | IVMP, TPE | 21 | EDSS NA  0.5, 0.7 | 2 | 0.3, 0.1 | NA | 0.5, 0.4 | vaccination | NA |
| Soni et al. | 10-14 | Female | NA | First attack | 0 | TM | IVMP,  TPE ×7 | 12 | EDSS 0  1.2, 1.2 | NA | 1.2, 1.2 | NA | NA | vaccination + penicillin | NA |

Abbreviations: Ecu, eculizumab; NA, Not available; EDSS, Expanded Disability Status Scale; VA, visual acuity; R, right eye; L, left eye; TM, transverse myelitis; IVMP, intravenous methylprednisolone pulse therapy; TPE, therapeutic plasma exchange; BS, brainstem; LON, left ON; RON, right ON; BON, both ON.

a, To protect patients’ privacy, precise ages were replaced.

**Figure 1** Vision deterioration before treatment in Patient 9
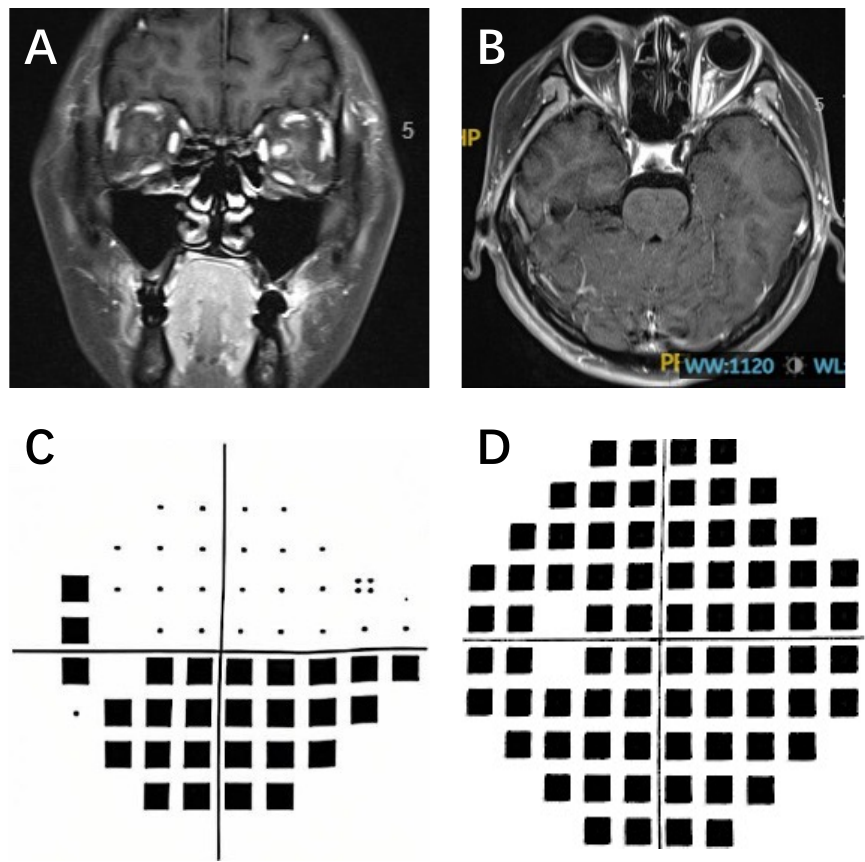

(A, B) MRI revealed a strikingly long-segment enhancement of the left optic nerve in the intraorbital segment, with pronounced nerve thickening and tortuosity. (C) The visual field (pattern deviation ) six days prior to eculizumab treatment; (D) The visual field on the day of eculizumab treatment(total deviation).
